# What Gets Funded Shapes What We Know: 15 Years of Canadian Women’s Health Research

**DOI:** 10.64898/2026.02.17.26346472

**Authors:** Laura L Gravelsins, Tallinn FL Splinter, Ahmad Mohammad, Samantha A Blankers, Gabrielle L Desilets, Liisa AM Galea

## Abstract

**Importance:** Funding of women’s health research has been low, with a narrow focus on what is considered women’s health. Understanding which lifespan stages and areas of women’s health are funded is essential to determine the breadth of women’s health research and identify where gaps in research are concentrated.

**Objective:** To examine which lifespan stages and areas of women’s health were more likely to be funded in open Canadian grant competitions.

**Evidence Review:** Publicly available funded Canadian Institutes of Health Research (CIHR) Project Grant abstracts from 2009 and 2023 were coded for mention of a hormonal transition period (puberty, menstrual cycle, pregnancy/postpartum, perimenopause/menopause), exogenous hormone use (hormonal contraception, fertility treatments, menopause hormone therapy), and/or a female-specific health condition. Abstracts were also coded for Indigenous health and Two Spirit, Lesbian, Gay, Bisexual, Trans, Transgender or Trans Identified, Queer, Intersex, Asexual, Plus (S2/LGBTQIA+) populations. Remaining grant abstracts were grouped by common theme.

Abstracts were analyzed for changes in research representation and funding over time and whether funding was lower than expected based on population prevalence or proportion of the lifespan spent in that stage.

**Findings:** Nearly 50% of female-specific research focused on cancers (breast, gynecologic) or pregnancy and did not significantly increase in funding or representation over time. Of the funded grant abstracts that focused on pregnancy, ∼22% examined outcomes pertaining only to the fetus/offspring, not the birthing parent. Over 15 years, 2.37% of *all CIHR* abstracts over 15 years were devoted to pregnancy, whereas only 0.24% was devoted to other hormonal life stages (menstrual cycles, menopause). For all hormonal transition stages except pregnancy, the proportion of grants and funding devoted to that stage was lower than expected based on the proportion of the lifespan spent in that stage.

**Conclusions and Relevance:** These findings reflect the narrow breadth of women’s health, which largely focused on cancers (breast, gynecologic) or pregnancy, rather than being distributed across key life course stages that shape women’s health. To advance science for all, the heterogeneity and complexity in women’s health across the lifespan must be embraced and barriers for women’s health research must be removed.

**Key Points:** *Question:* Which areas and life stages of women’s health are most likely to be funded in Canadian open grant competitions, and where are funding gaps concentrated?

*Findings:* Nearly half of female-specific grants focused on cancer or pregnancy, with little change over time. Pregnancy dominated hormonal-stage research, often excluding maternal outcomes, while menstrual and menopausal stages were rarely funded. For most life stages, funding was lower than expected based on lifespan representation.

*Meaning:* Women’s health research funding remains narrowly focused. Broader, life-course–inclusive investment is needed to address critical gaps and advance equitable health science.

## Introduction

Women’s health has historically been undervalued and underfunded^1^ which in part led to mandates to incorporate sex or gender into research projects by major funding agencies^2^. Yet, women’s health continues to be unprioritized^3,4^, resulting in fewer publications on women’s health^5^. The breadth of women’s health research is important to recognise, as women’s health goes beyond disorders that only affect women, to those that disproportionately affect women, or that show differences in manifestation or treatment efficacy. Thus it is essential to understand how the unique characteristics of female biology and the intersection of the psychosocial world can differentially impact health for women. Women’s health is more than reproductive health^6^ and diseases that disproportionately influence females are underfunded in North America^1,4^. Here we examined what lifespan stages and areas of women’s health were more likely to be funded in part to determine where gaps in research are concentrated for women’s health. We expected that more funding would be dedicated to women’s reproductive health, including pregnancy and fertility.

## Methods

We aimed to investigate the scope of women’s health research over 15 years, from 2009-2023 examining the major funding mechanism in Canada for investigator driven research: Canadian Institutes for Health Research (CIHR) operating/project grants (akin to RO1 at the National Institute of Health (NIH)). Using methods consistent with our previous analyses^4,7^, we categorized publically available grant abstracts as female-specific (i.e., mentioning studying females or women only) and coded whether or not abstracts mentioned investigating a particular hormonal transition period (puberty, menstrual cycle, pregnancy/postpartum, perimenopause/menopause), exogenous hormone use (hormonal contraception, fertility treatments, menopause hormone therapy), and/or a female-specific health condition (e.g., endometriosis, polycystic ovary syndrome, catamenial epilepsy, Turner Syndrome). We also coded abstracts that identified Indigenous health, and Two Spirit, Lesbian, Gay, Bisexual, Trans, Transgender or Trans Identified, Queer, Intersex, Asexual, Plus (S2/LGBTQIA+). Remaining grant abstracts were grouped by common theme, or coded as “Other” if they could not be grouped.

Linear regressions were conducted to investigate if the percentage of grant abstracts and funding dollars dedicated to these studied areas of female health increased over time with a false discovery rate (FDR) correction for multiple comparisons and statistical significance set to *p* < 0.05. Partial eta-squared was calculated as a measure of effect size. To evaluate if a particular hormonal transition, exogenous hormone use, or female-specific health condition received less focus or funding than would be expected based on the prevalence within the population, we calculated the representation fold difference and funding fold difference.

## Results

Over 15 years, there were 681 funded female-specific grants that received $441.18 million in research funding. More than 50% of funded female-specific grant abstracts were devoted to pregnancy or cancer (gynecological or breast) (Fig 1A) and collectively received approximately 70% of research funding that was devoted to female-specific grants ($307.88 million). However, ∼22% of the funded grant abstracts that focused on pregnancy, examined outcomes pertaining only to the fetus/offspring (Fig 2A), and not the birthing parent. This includes abstracts which examined how maternal conditions (e.g., distress, substance use, or disease) affected the developmental outcomes of the fetus/offspring, but did not mention maternal outcomes. The percentage of funded grants and amount of funding devoted to these areas of women’s health did not significantly increase over 15 years (Fig 1B; Supplementary Material Table 1).

**Table 1.** Alignment of CIHR Funding With Female Life Course and Population Health Burden. For lifecourse stage domain, the “% Reference Exposure” refers to the percentage of the female lifespan spent in that life course stage and was calculated assuming the Canadian female average life expectancy from birth of 83.9 years old^9^. The “Fold Difference” was calculated by dividing the “% of Female-Specific CIHR Grants” by the “% Reference Exposure”, such that a value of 1 indicates the amount of representation/funding or is proportional to the life span exposure, whereas a value greater than 1 indicates over representation/over funding and a value less than 1 indicates underrepresentation/underfunding. Two menopause grant abstracts examined both the perimenopausal and postmenopausal periods, and one grant abstract examined predictors of early menopause and was therefore coded as neither perimenopause nor postmenopause. Eleven of the contraception-focused grant abstracts examined hormonal contraceptives (HC) specifically.

| Domain | Category | Duration (% of Lifespan) | % Reference Exposure* | Metric | % All CIHR Grants | % Female-Specific CIHR Grants | Fold Difference† |
| --- | --- | --- | --- | --- | --- | --- | --- |
| Life Course stage | Puberty | 2–5 years | 2.38–5.96 | Representation | 0.04 | 0.59 | 0.10–0.25 |
|  |  |  |  | Funding | 0.02 | 0.34 | 0.06–0.14 |
|  | Pregnancy | 0.94 years (average of 1.25 pregnancies) | 1.12 | Representation | 2.37 | 37 | 33 |
|  |  |  |  | Funding | 2.25 | 35.3 | 31.52 |
|  | Pre-menopause (menstrual cycle/cycling) | ~38.28 years | 45.63 | Representation | 0.03 | 0.44 | 0.01 |
|  |  |  |  | Funding | 0.02 | 0.31 | 0.007 |
|  | Perimenopause (n=4) | ~2–10 years | 2.38–11.92 | Representation | 0.04 | 0.59 | 0.05–0.21 |
|  |  |  |  | Funding | 0.02 | 0.34 | 0.03–0.14 |
|  | Post-menopause (n=16) | ~32.9 years | 39.21 | Representation | 0.15 | 2.35 | 0.06 |
|  |  |  |  | Funding | 0.15 | 2.45 | 0.06 |
| Area of Female Health | Breast Cancer | -- | 25.7 | Representation | 1.23 | 19.24 | 0.75 |
|  |  |  |  | Funding | 1.38 | 21.64 | 0.84 |
|  | Gynecologic Cancers | -- | Ovary: 2.5; Cervix: 1.3; Uterus: 7.0 | Representation | 0.97 | 15.12 | 1.4 |
|  |  |  |  | Funding | 1.01 | 15.81 | 1.46 |
|  | Lung Cancer | -- | 14.2 | Representation | 0.02 | 0.29 | 0.02 |
|  |  |  |  | Funding | 0.03 | 0.41 | 0.03 |
|  | Endometriosis | -- | 10 | Representation | 0.15 | 2.35 | 0.24 |
|  |  |  |  | Funding | 0.13 | 2.1 | 0.21 |
|  | PCOS | -- | 6.71 | Representation | 0.06 | 0.88 | 0.13 |
|  |  |  |  | Funding | 0.03 | 0.55 | 0.08 |
|  | Hormonal Contraception (n=11) | -- | 29 | Representation | 0.1 | 1.62 | 0.06 |
|  |  |  |  | Funding | 0.06 | 0.96 | 0.03 |
|  | Menopausal Hormone Therapy | -- | 9.5 | Representation | 0.04 | 0.59 | 0.06 |
|  |  |  |  | Funding | 0.03 | 0.51 | 0.05 |
|  | Indigenous Women's Health | -- | 8.67 | Representation | 0.28 | 4.41 | 0.51 |
|  |  |  |  | Funding | 0.36 | 5.69 | 0.66 |
|  | 2S/LGBTQIA+ | -- | 4.4 | Representation | 0.08 | 1.32 | 0.3 |
|  |  |  |  | Funding | 0.11 | 1.73 | 0.39 |
|  | Sexual Health Conditions | -- | ~39 | Representation | 0.71 | 11.01 | 0.28 |
|  |  |  |  | Funding | 0.72 | 11.3 | 0.29 |
|  | Fertility | -- | 15.7 | Representation | 0.28 | 4.41 | 0.28 |
|  |  |  |  | Funding | 0.29 | 4.55 | 0.29 |
|  | Sexual Assault / Domestic Violence | -- | 33.3 | Representation | 0.16 | 2.5 | 0.08 |
|  |  |  |  | Funding | 0.17 | 2.69 | 0.08 |

**Fig 1.**
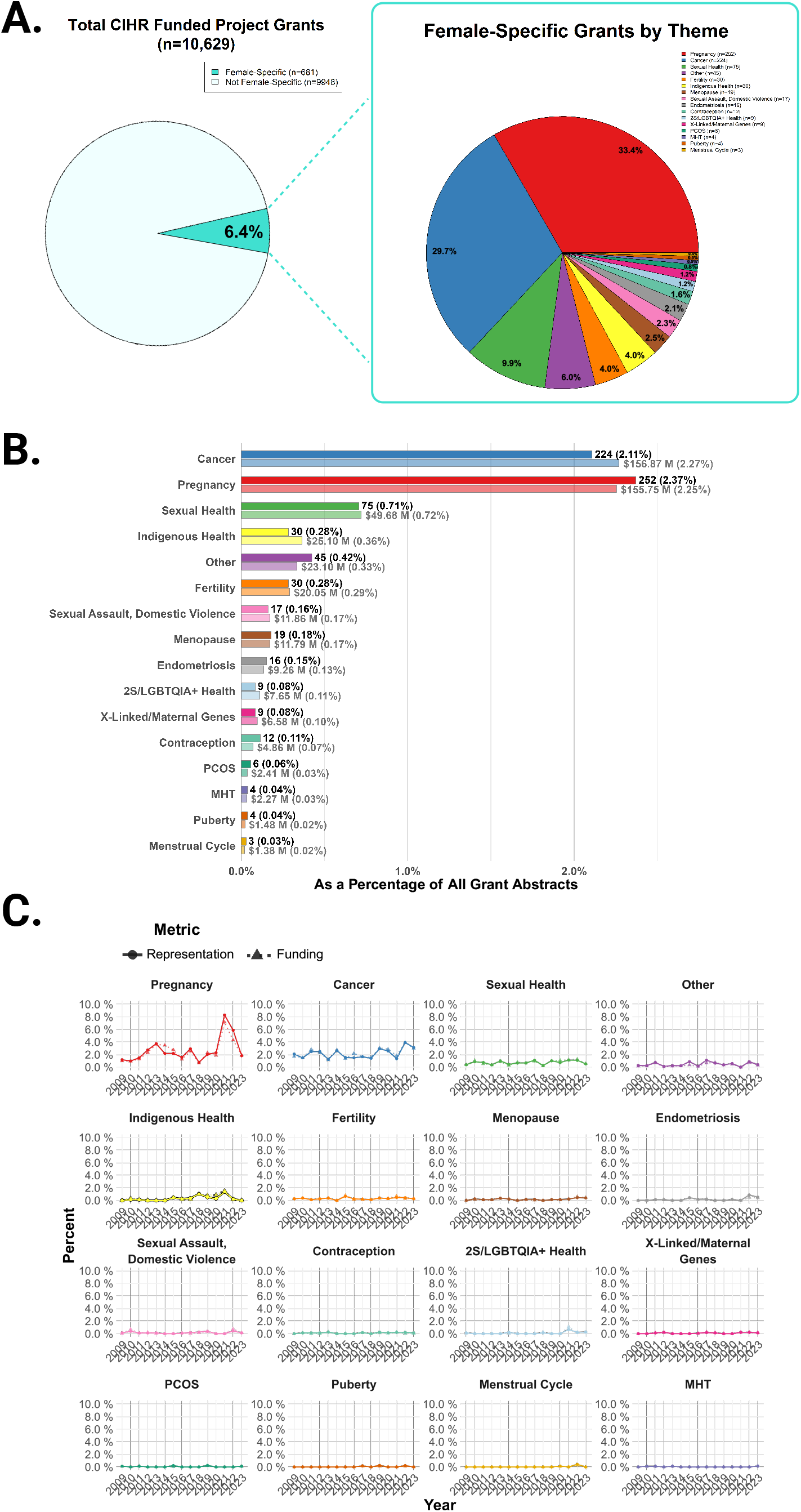
A) Breakdown of the 10,629 female-specific grant abstracts over 15 years. Note 67 grant abstracts were coded in two or more categories, e.g., all grants coded in “Menopause hormone therapy (MHT)” were also coded in “Menopause”, so the denominator used to generate the pie chart is 755 to reflect the distribution of category assignments (i.e., 681 coded once + 62 coded twice + 4 coded three times + 1 coded 5 times). “Sexual assault, domestic violence” includes childhood maltreatment. “Pregnancy” includes the perinatal and postpartum periods (n=248) as well as grants on parenting or mother-child dyad (n = 4). “Sexual health” includes sexual function, genito-pelvic pain, vestibulodynia, sexual distress, vaginismus, low sexual desire, urinary incontinence, pelvic floor disorders, and sexually transmitted infections. “Contraception” includes both hormonal and non-hormonal contraception. “Fertility” includes assisted fertility treatments and technologies, and biological mechanisms supporting reproductive success. B)Breakdown of the 681 female-specific grant abstracts from 2009-2023 according to area of research by representation (darker bars) and funding (lighter bars) as a proportion of all grant abstracts. Funding amounts are provided in millions (M). C) Amount of representation and funding devoted to areas of women’s health over time from 2009-2023. Breakdown is provided as a proportion of all funded CIHR grant abstracts. Research representation is represented by circles and research funding is represented by triangles. PCOS = polycystic ovary syndrome; 2S/LGBTQIA+ = Two-Spirit, Lesbian, Gay, Transgender, Queer, Intersex, Asexual, Plus; MHT = Menopause hormone therapy.

**Figure 2.**
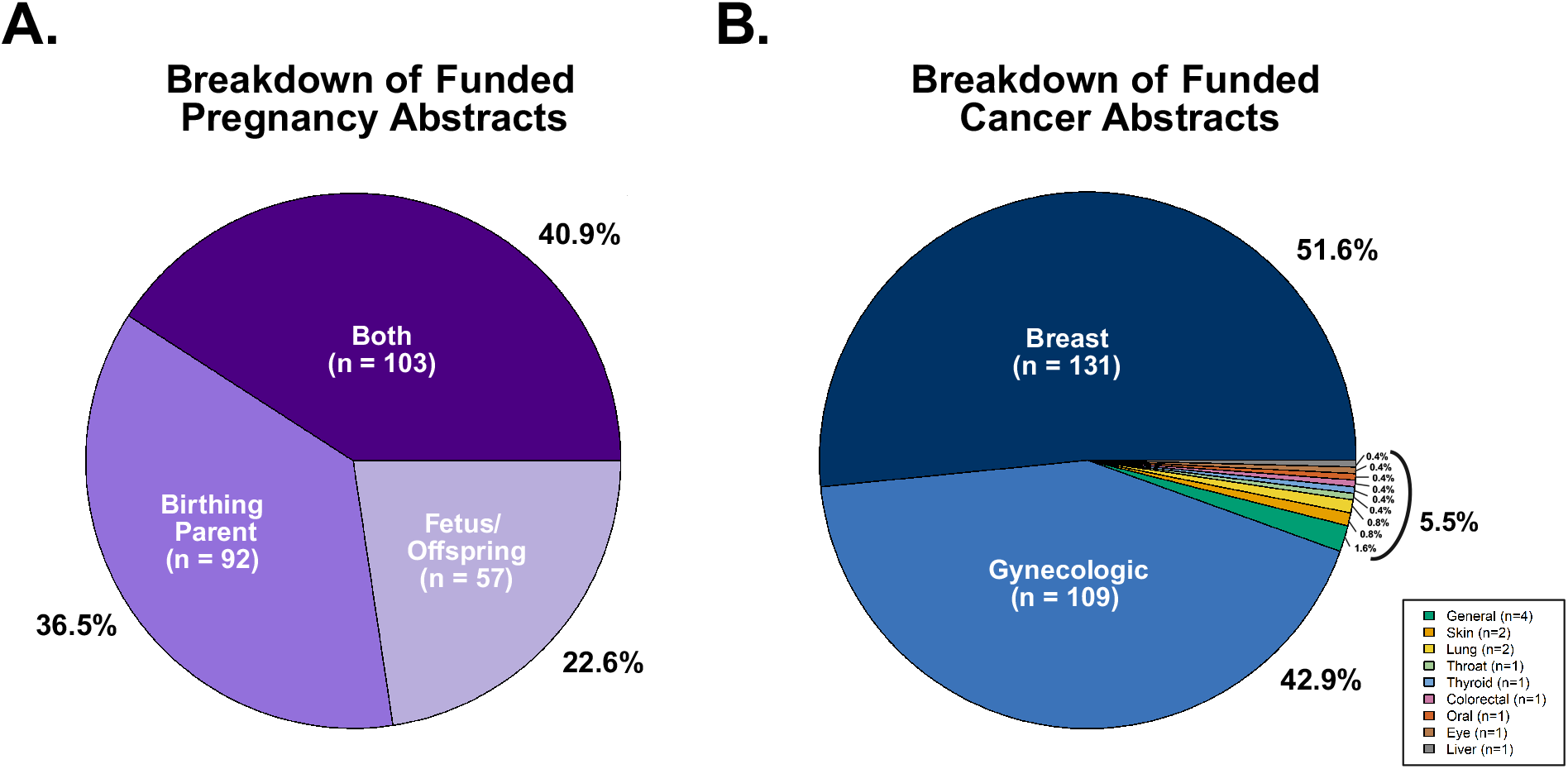
A) Breakdown of funded pregnancy abstracts by focus on the birthing parent, fetus/offspring, or both. B) breakdown of funded cancer abstracts by cancer type. Note: Gynecologic cancers include ovarian, cervical, endometrial, fallopian tube, vulvar, and uterine cancers. Twenty-three cancer abstracts were coded in more than one type of cancer (17 coded twice, 5 coded three times, 1 coded 4 times), so the denominator used is 254.

Compared to all other hormonal transition stages, there was a disproportionate focus on pregnancy. While 37.0% (n=252) of *all female-specific* abstracts (n=681) were devoted to pregnancy, only 2.8% (n=19) were devoted to menopause, 0.44% (n=3) to the reproductive period/menstrual cycle, and 0.59% (n=4) to puberty. In other words, 2.37% (n=252) of *all CIHR* abstracts over 15 years (n=10,629) were devoted to pregnancy, whereas only 0.24% was devoted to other hormonal life stages (0.18% (n=19) were devoted to menopause, 0.03% (n=3) to the reproductive period/menstrual cycle, and 0.04% (n=4) to puberty) (Fig 1B). Similar trends were observed in funding (Table 1). This gap is even more apparent when you factor in that females spend ∼45.6% of their lifespan with menstrual cycles, and ∼39.2% of their lifespan in menopause, but will spend only ∼1.1% of their lifespan in pregnancy (Table 1). For all hormonal transition stages except pregnancy, the proportion of grants and funding devoted to that stage was lower than expected based on the proportion of the lifespan spent in that stage (Table 1).

Of all funded cancer abstracts in females, 94.5% focused on breast or gynecologic cancers (e.g., ovarian, endometrial, vulvar, cervical; Fig 2B). Based on the prevalence of breast and gynecologic cancers in the Canadian population, it is clear that these areas received a disproportionate amount of focus (Table 1). In contrast, lung cancer accounted for 0.8% of funded cancer grants, but is the second leading cause of cancer in women accounting for 14.2% of cases^8^.

## Discussion

Our results suggest that the breadth of funded women’s health research in Canada across 15 years is limited in scope. Nearly two-thirds of grant abstracts were devoted to gynecologic/breast cancer and pregnancy, and collectively received the vast majority of women’s health research funding (∼70%). Furthermore, ∼20% of the pregnancy grants were focused on offspring/child outcomes rather than the pregnant individual, further reflecting the devaluation of women’s health.

Research funding across the female lifespan is skewed toward pregnancy rather than distributed across life stages. On average, females spend 1% of their lifespan pregnant yet this lifestage received the majority of women’s health funding. In contrast, females spend ∼40% of their lifespan in menopause which accounts for only 2.73% of all funded female-specific grants. To advance science, the heterogeneity and complexity in women’s health across the lifespan must be embraced.

Menopause is a biologically significant aging inflection point in females, and a key intervention point for healthy aging. Yet over 15 years, only 19 of all funded CIHR grant abstracts mentioned menopause (0.18%; 19 of 10,629), 4 of which investigated the effects of MHT (0.04%; 4 of 10,629), which has been associated with increased quality of life and reduced dementia risk in those over 65^10^. Indeed, women in their 60s are twice more likely to be diagnosed with Alzheimer’s Disease compared to breast cancer^11^, reflecting the growing need for more dedicated aging health research in women.

Menstrual cycles are considered a barometer of current and future health in menstruating individuals, including a potential indicator of later-life cardiovascular disease^12^. Menstruation increases risk for migraines, epileptic seizures, and autoimmune flare-ups, significantly influencing quality of life^13,14^. Endometriosis, PCOS, and PMDD are chronic conditions linked to the menstrual cycle, yet also remain understudied. To put things in perspective, menstruating individuals will experience ∼451 menstrual cycles over their lifetime or ∼3,500 days menstruating, corresponding to ∼34.7 years of menstrual activity^15^. Yet our analyses revealed that only 0.31% of all female-specific funded grants (0.02% across all grants) were focussed on menstrual cycles.

Puberty is associated with an increased risk for mood and anxiety disorders in females^16^. Hormonal contraceptives (HC) are used by ∼80% of the population and approximately 300 million people globally^17^. HCs can reduce risk of endometrial and ovarian cancer, but can also increase depression risk, depending on age and formulation^18,19^. Yet again grant funding remained low with funded abstracts mentioning HC or puberty collectively receiving 0.04% of all CIHR funding.

Studying women’s health issues will not just be informative for women and assigned female at birth (AFAB), it will also help other genders. It can identify and refine sex/gender-specific diagnostic criteria that may be needed to correct for delays in diagnosis^20^. Moreover, hormones such as estrogens and progesterone, are not female-specific and a better characterization of their biological influences will translate to all sexes and genders. Knowledge gained from studying MHT will inform on continued gender affirming hormone therapy use in older, transgender individuals.

Although, our findings indicate that in women’s health, more grants were funded on pregnancy and gynecologic/breast cancers, these areas of research were still underfunded compared to the population affected. It is necessary to better understand which stage of the funding cycle poses barriers for women’s health research: is it a lack of researchers, a lack of awareness of what constitutes women’s health, and/or do reviewers undervalue the importance of women’s health research? Large discoveries have been made in women’s health that are beyond reproductive health, leading to promising new therapeutics including brexanalone, a synthetic allopregnanlone and treatment for postpartum depression^21^; cellular communication network factor 3 (CCN3), a new bone hormone and possible treatment for bone loss^22^; and growth differentiation factor 15 (GDF15) as the mechanism for severe nausea and vomiting during pregnancy^23^. These discoveries have implications for treatments in men and other genders across disorders. Economic analyses suggest that a trillion dollars a year could be saved if we close the gap in women’s health^24^. Bridging this gap necessitates a strategic commitment to research and our analyses suggest the current landscape of funded research in women has a narrow focus. Finally, international surveys converge on a clear message: women themselves prioritise menopausal hormone health and mental health as critical, yet under-researched, areas of their health^25,26^. In the context of clear public support, policy-makers have both the mandate and opportunity to strategically invest in a broader, lifespan-inclusive women’s health research agenda.

## Data Availability

All data produced in the present study are available upon reasonable request to the authors

## Authors’ Contributions

L.L.G. and L.A.M.G. designed the study and were responsible for the methodology. L.L.G. was responsible for formal analysis, data visualisation, and wrote the original draft of the manuscript. L.L.G., T.F.L.S., A.M., S.A.B., and G.L.D. contributed to data curation, including data coding. L.A.M.G. was responsible for funding acquisition and study supervision. All authors reviewed and edited the manuscript.

## Acknowledgements

We are deeply grateful to Tori N. Stranges and Amanda B. Namchuk for their invaluable contributions in coding abstracts from 2009-2020.

## Supplementary Material

**Table 1.** Linear Model Output Showing Funding and Representation Does Not Significantly Increase Over Time.

| Data Type | Area of Women's Health | Estimate | SE | t-statistic | p-value | Adjusted p-value | $\eta^2$ |
| --- | --- | --- | --- | --- | --- | --- | --- |
| Funding | Cancer | -173.15 | 85.90 | -2.02 | 0.07 | 0.27 | 0.24 |
| Funding | Pregnancy | -316.04 | 189.59 | -1.67 | 0.12 | 0.27 | 0.18 |
| Funding | Sexual health | -64.78 | 38.35 | -1.69 | 0.12 | 0.27 | 0.18 |
| Funding | Indigenous health | -95.89 | 51.76 | -1.85 | 0.09 | 0.27 | 0.21 |
| Funding | Other | -32.35 | 30.77 | -1.05 | 0.31 | 0.45 | 0.08 |
| Funding | Fertility | -18.48 | 24.94 | -0.74 | 0.47 | 0.60 | 0.04 |
| Funding | Sexual Assault / Domestic Violence | -4.72 | 30.11 | -0.16 | 0.88 | 0.92 | 0.00 |
| Funding | Menopause | -39.64 | 19.00 | -2.09 | 0.06 | 0.27 | 0.25 |
| Funding | Endometriosis | -35.79 | 18.03 | -1.98 | 0.07 | 0.27 | 0.23 |
| Funding | 2S/LGBTQIA+ Health | -58.56 | 31.29 | -1.87 | 0.08 | 0.27 | 0.21 |
| Funding | X-Linked / Maternal Genes | -22.66 | 11.55 | -1.96 | 0.07 | 0.27 | 0.23 |
| Funding | Contraception | -3.96 | 10.86 | -0.36 | 0.72 | 0.82 | 0.01 |
| Funding | PCOS | 3.42 | 6.82 | 0.50 | 0.63 | 0.74 | 0.02 |
| Funding | MHT | -0.91 | 6.68 | -0.14 | 0.89 | 0.92 | 0.00 |
| Funding | Puberty | -8.47 | 5.77 | -1.47 | 0.17 | 0.29 | 0.14 |
| Funding | Menstrual Cycle | -14.52 | 8.57 | -1.69 | 0.11 | 0.27 | 0.18 |
| Representation | Cancer | -138.03 | 91.12 | -1.51 | 0.15 | 0.29 | 0.15 |
| Representation | Pregnancy | -422.42 | 221.07 | -1.91 | 0.08 | 0.27 | 0.22 |
| Representation | Sexual Health | -40.89 | 34.35 | -1.19 | 0.26 | 0.39 | 0.10 |
| Representation | Indigenous Health | -81.91 | 44.31 | -1.85 | 0.09 | 0.27 | 0.21 |
| Representation | Other | -30.07 | 39.59 | -0.76 | 0.46 | 0.60 | 0.04 |
| Representation | Fertility | -13.96 | 19.61 | -0.71 | 0.49 | 0.60 | 0.04 |
| Representation | Sexual Assault / Domestic Violence | -5.32 | 17.95 | -0.30 | 0.77 | 0.85 | 0.01 |
| Representation | Menopause | -24.80 | 15.50 | -1.60 | 0.13 | 0.28 | 0.17 |
| Representation | Endometriosis | -58.01 | 27.16 | -2.14 | 0.05 | 0.27 | 0.26 |
| Representation | 2S/LGBTQIA+ Health | -38.77 | 20.21 | -1.92 | 0.08 | 0.27 | 0.22 |
| Representation | X-Linked / Maternal Genes | -15.54 | 11.16 | -1.39 | 0.19 | 0.30 | 0.13 |
| Representation | Contraception | -16.23 | 11.15 | -1.46 | 0.17 | 0.29 | 0.14 |
| Representation | PCOS | 0.56 | 11.13 | 0.05 | 0.96 | 0.96 | 0.00 |
| Representation | MHT | 5.58 | 6.85 | 0.81 | 0.43 | 0.60 | 0.05 |
| Representation | Puberty | -16.16 | 10.41 | -1.55 | 0.14 | 0.29 | 0.16 |
| Representation | Menstrual Cycle | -22.57 | 12.84 | -1.76 | 0.10 | 0.27 | 0.19 |

